# Multi-omics empowered deep phenotyping of ulcerative colitis

**DOI:** 10.1101/2022.05.25.22275502

**Authors:** Lukas Janker, Dina Schuster, Patricia Bortel, Gerhard Hagn, Julia Brunmair, Samuel M. Meier-Menches, Johanna C. Mader, Astrid Slany, Andrea Bileck, Christian Madl, Lukas Unger, Barbara Hennlich, Barbara Weitmayr, Giorgia Del Favero, Dietmar Pils, Tobias Pukrop, Nikolaus Pfisterer, Thomas Feichtenschlager, Christopher Gerner

## Abstract

**Objective:** Ulcerative colitis (UC) is a chronic disease with rising incidence and unclear etiology. The application of mass spectrometry-based analysis methods shall support the establishment of systemic molecular biomarker signatures providing status information with regard to individual UC pathomechanisms.

**Design:** UC pathomechanisms were assessed by proteome profiling of human tissue specimen, obtained from five distinct colon locations each of 12 patients. Systemic disease-associated alterations were investigated in a cross-sectional setting by mass spectrometry-based multi-omics analyses comprising proteins, metabolites and eicosanoids of plasma obtained from UC patients during disease and upon remission in comparison to healthy controls.

**Results:** Tissue proteome profiling identified colitis-associated activation of neutrophils, macrophages, B- and T-cells, fibroblasts, endothelial cells and platelets, and indicated hypoxic stress, as well as a general downregulation of mitochondrial proteins accompanying the establishment of apparent wound healing-promoting activities including scar formation. While the immune cells mainly contributed pro-inflammatory proteins, the colitis-associated epithelial cells, fibroblasts, endothelial cells and platelets predominantly formed anti-inflammatory and wound healing-promoting proteins. Blood plasma proteomics indicated chronic inflammation and platelet activation, whereas plasma metabolomics identified disease-associated deregulation of bile acids, eicosanoids and gut microbiome-derived metabolites. Upon remission, several, but not all, molecular candidate biomarker levels recovered to normal levels. These findings may indicate that pathomechanisms related to gut functions, gut microbiome status, microvascular damage and metabolic dysregulation associated with hypoxia may not resolve uniformly during remission.

**Conclusions:** This study integrates and expands the knowledge about local and systemic effects of UC and identifies biomarker profiles related to molecular UC pathomechanisms.

## Introduction

Ulcerative colitis (UC), an inflammatory bowel disease (IBD), is characterized by ascending inflammation of the large intestine with intermittent cycles of active inflammation and asymptomatic periods.^1^ With an early disease onset at around 30 years of age and increasing incidence rates in developed countries (24.3/100,000 in Europe; 19.2/100,000 in North America), UC is becoming an increasing and severe health risk for millions of people.^2, 3^ Due to the chronic inflammation, UC patients suffer from an increased risk of colorectal cancer as well as thromboembolic complications.^4-6^ The chronicity of UC accompanied with life-long non-curative treatment of symptoms is challenging, as patients are treated based on symptom subsets and their severity in an iterative scheme^1, 7-10^, which has not fundamentally changed for the past two decades.^5, 11, 12^ Often used metrics including the Ulcerative Colitis Disease Activity Index (UCDAI) and MAYO-score are reliant on multiple clinical observations, invasive and non-invasive in nature, as well as on the compliance of patients subjective measures in form of patient-reported outcomes, so called PRO reports.^13, 14^ A vast effort was put into the elucidation of disease driving genetic factors including genome-wide association studies, but to the best of our knowledge, no definitive causal links could be established yet.^15^ MHC locus HLA Class II alleles as well as the multidrug resistance gene *MDR1* have been implicated as possible genetic susceptibility factors.^16-18^ Studies focusing on the genetic landscape of the second main type of IBD, namely Crohn’s Disease (CD), found, in contrast to UC, evidence of causal effects of single nucleotide polymorphisms with biological plausibility and dose-response effects independently verified by multiple studies.^19-22^ The abovementioned points strengthen the theory of involvement of environmental factors and other possible post-genomic related influence factors such as the general composition and functional state of the proteome, metabolome and microbiome. Even for malignant diseases where genetic instability determines disease progression, post-genomic analysis tools can provide deeper insights into molecular mechanisms of progression and escalation.^23^ Recent studies^24, 25^ provide insights into potential pathophysiologic mechanisms of UC, showing for example the influence of the microbiome and its compositional alterations before and after diagnosis with UC^26^, the activation of the innate immune system monitored on a proteome wide scale^27^, or general compositional changes in the metabolome^28, 29^ or microbiome dependent co-metabolome.^30, 31^ Although these findings are leading UC related research into a more molecularly focused direction, there are still pathophysiological processes, which remain to be unveiled. Studies focusing on *post hoc* analysis of serological markers for the discrimination of clinically relevant cohorts showed the limitations of established marker molecules, and demonstrated the need for comprehensive screening and combination of data on multiple “omics” levels.^32, 33^ In the presented study, we thus aimed to investigate pathophysiological aberrations *in situ*. We present an in-depth analysis of intra-individual colonic proteome alterations during active UC in a spatially resolved manner. The parallel analysis of plasma samples, allowing to identify systemic disease-associated alterations, was performed by the combination of an extensive panel of biomolecules ranging from amino acids to lipids, including eicosanoids and plasma proteins. An additional case-control study with UC patients in remission was performed in order to investigate remission-associated normalization of biomarker profiles. Thus, we demonstrate that specific colitis-associated biomarkers were still deregulated at a time when no more symptoms were reported and those indicate possible determinants for chronicity. The present multi-omics analyses provide a deep molecular phenotyping of UC disease manifestation, pointing at novel therapeutic targets and presenting biomarker candidates that may be validated in prospective clinical trials.

## Material and Methods

A more detailed description of the methodology can be found in the Supplementary Materials and Methods.

### Study design and population

The presented case-control study includes patients from the age of 18 and above diagnosed with ulcerative colitis (UC), either in active or remission state, as well as healthy control patients, as described in Figure 1A. Patient information regarding clinical data from active and remission patients, as well as age distribution in regard to healthy controls can be found in Supplementary Tables S1, S2 and Suppl. Figure S1. Exclusion criteria for active UC patients included pancolitis, remission status according to MAYO-Score, infections, colon resection or colectomy as well as colitis indeterminata. Exclusion criteria for remission UC patients included infections, colitis indeterminata as well as colon resection or colectomy. Patients with active UC underwent a routine colonoscopy for the assessment of the local inflammatory status including fecal calprotectin and assignment of MAYO-Score.^13^ Colon biopsies were classified into three categories ranging from non-inflamed over surrounding/mildly inflamed to inflamed. Classification was based on histological findings of inflammatory features (Supplementary Figure S2. For the monitoring of systemic events happening during the pathogenesis of UC, blood plasma was collected from each patient cohort. The study was approved by the ethics commission of the city of Vienna with votum EK 18-193-0918. All patients gave informed consent.

**Figure 1:**
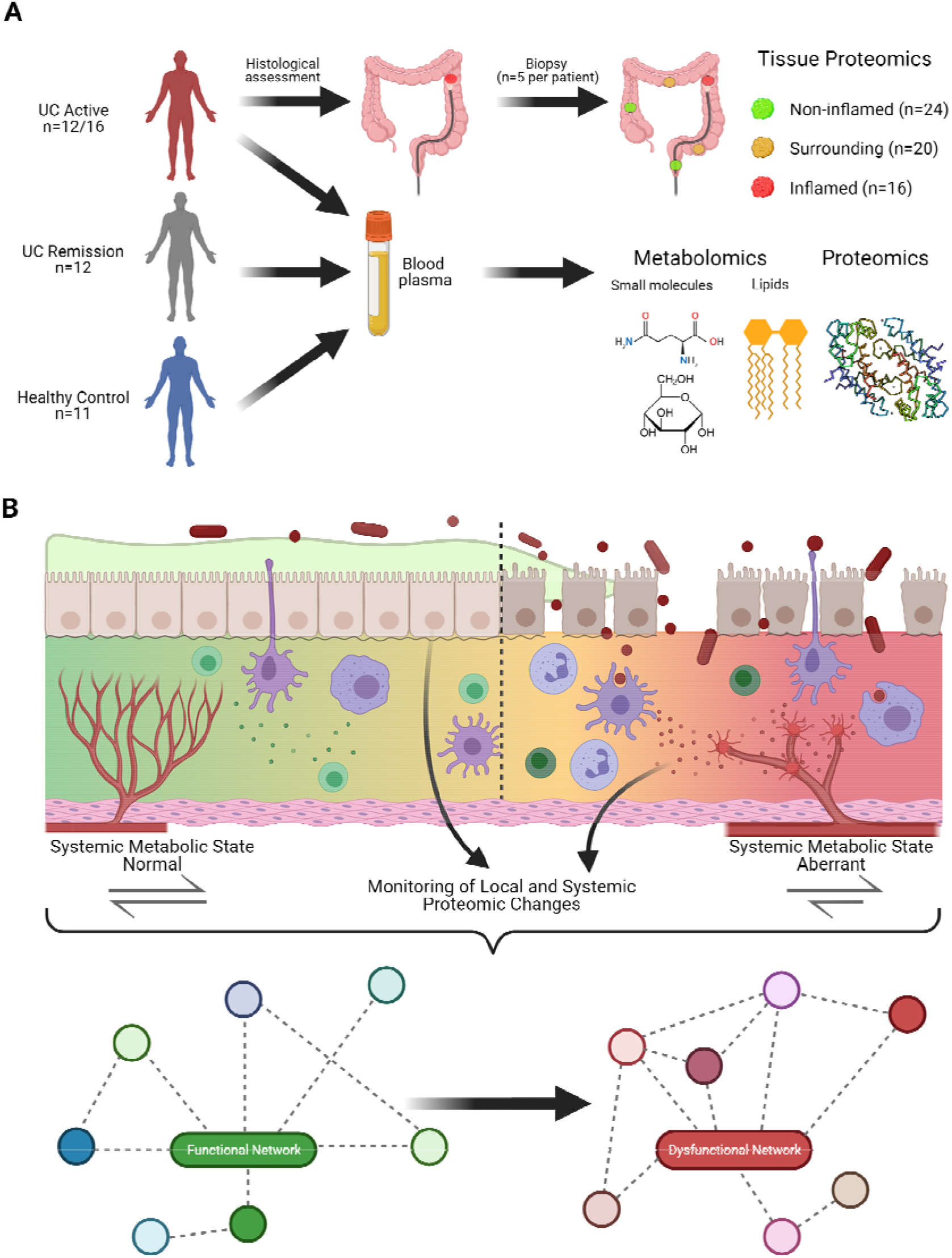
Study design supporting the discovery of functional biomarker. **(A)** Patient cohort and general sampling strategy. Patients with active UC (top, n=12 for complete tissue and plasma series, n=4 only plasma available) providing tissue (n=5 per patient) and plasma samples. Blood plasma samples from UC patients in remission (n=12), as well as healthy controls (n=11) were collected. Based on histological classification, tissue samples were categorized into non-inflamed (n=24), surrounding (n=20) and inflamed (n=16) tissue. **(B)** Schematic overview regarding pathophysiological processes taking place in different compartments. Blood microvessels supply the mesenchymal interstitium separated from the mucus-forming parenchyme via a basal membrane. Disease progression (left to right) associated with microvascular damage, invasion of immune cells, breakdown of the basal membrane, loss of epithelial cells and increased access of inflammation-inducing bacteria are depicted, together with the presently applied complementary analysis strategies.

### Sample collection and processing

Colon biopsies of colonic subsections from active UC patients were taken, washed with PBS and immediately frozen at −80°C or processed for subsequent histologic examination. Blood was drawn from all three cohorts, processed into blood plasma and frozen following a strict time regimen. A detailed description of the methodology can be found in the Supplementary Materials and Methods.

### Statistical data analysis

Proteomics data was analyzed with MaxQuant (version 1.6.17.0).^34^ Eicosanoid spectra were compared with reference spectra from the Lipid Maps depository library and subsequently integrated using the TraceFinderTM software package (version 4.1).^35^ Targeted metabolomics data validation and evaluation was performed with the software supplied with the MxP® Quant 500 Kit (MetIDQ-Oxygen-DB110-3005). For the analysis of causal networks between inflamed and non-inflamed tissue, the software CausalPath with standard parameter settings and pre-processed LFQ intensity values (Supplementary Table S3) was utilized.^36^ Additional statistical analysis was performed utilizing Perseus (version 1.6.14.0), Microsoft Excel and GraphPad Prism (version 6.07).

Omics data were integrated in R using N-integration discriminant analysis with DIABLO (Data Integration Analysis for Biomarker discovery, R-package mixOmics 6.18.0,^37^) and an own developed method using Gaussian Graphs Models Selection (R-package GGMselect 0.1-12.4, CIT2) for analytes reduction via sub-network generation and Principal Component representation. Subsequently partial correlations of significantly deregulated single analytes with analyte sub-network representations were performed.^38^

## Results

### Rationale and presentation of the study design

After informed consent, five colon biopsy samples from different locations were obtained from each of the 12 patients suffering from active ulcerative colitis (clinical parameters are provided in Supplementary data). The tissue samples were categorized via histological findings into “inflamed”, “surrounding” and “non-inflamed” (Figure 1A, Suppl. Figure S1). Blood plasma was collected from a total of 16 acute UC patients, including the 12 patients that underwent biopsy, as well as other 12 UC patients after successful remission (clinical parameters are provided in Supplementary data and Supplementary Tables S7, S8). Blood plasma from 11 healthy donors served as controls. Each plasma sample was analyzed with regard to proteins, metabolites and eicosanoids using three different mass spectrometry-based analysis methods (Figure 1A). Tissue proteome analysis was conducted to provide insight into disease-associated pathomechanisms, and plasma samples from the same patients were analyzed to investigate whether local changes were indicated by systemic plasma profiles (Figure 1B).

### Tissue proteomics reveals molecular patterns associated with local UC pathomechanisms

The analysis of a total of 60 tissue biopsy samples resulted in the identification of 4,579 proteins (at least two peptides per protein, FDR < 0.01 at protein and peptide level, at least 5 independent identifications in at least one tissue category per protein, Supplementary Table S3) and various protein regulatory events distinguishing the three pre-defined tissue categories (Figure 2A, Supplementary Table S4). PCA separated “inflamed” versus “non-inflamed”, while the surrounding tissue samples were found dispersed across the other two groups (Figure 2B). It is fair to expect alterations of the cell type composition when comparing such tissue categories. Thus, differentially regulated proteins (FDR < 0.01) were attributed in a first step to different cell types according to expression specificity. In a subsequent step, functional cell activation markers were considered as described previously,^39-41^ in addition to protein regulatory events pointing to characteristic pathomechanisms (Figure 2C, Supplementary data, Supplementary Table S5).

**Figure 2:**
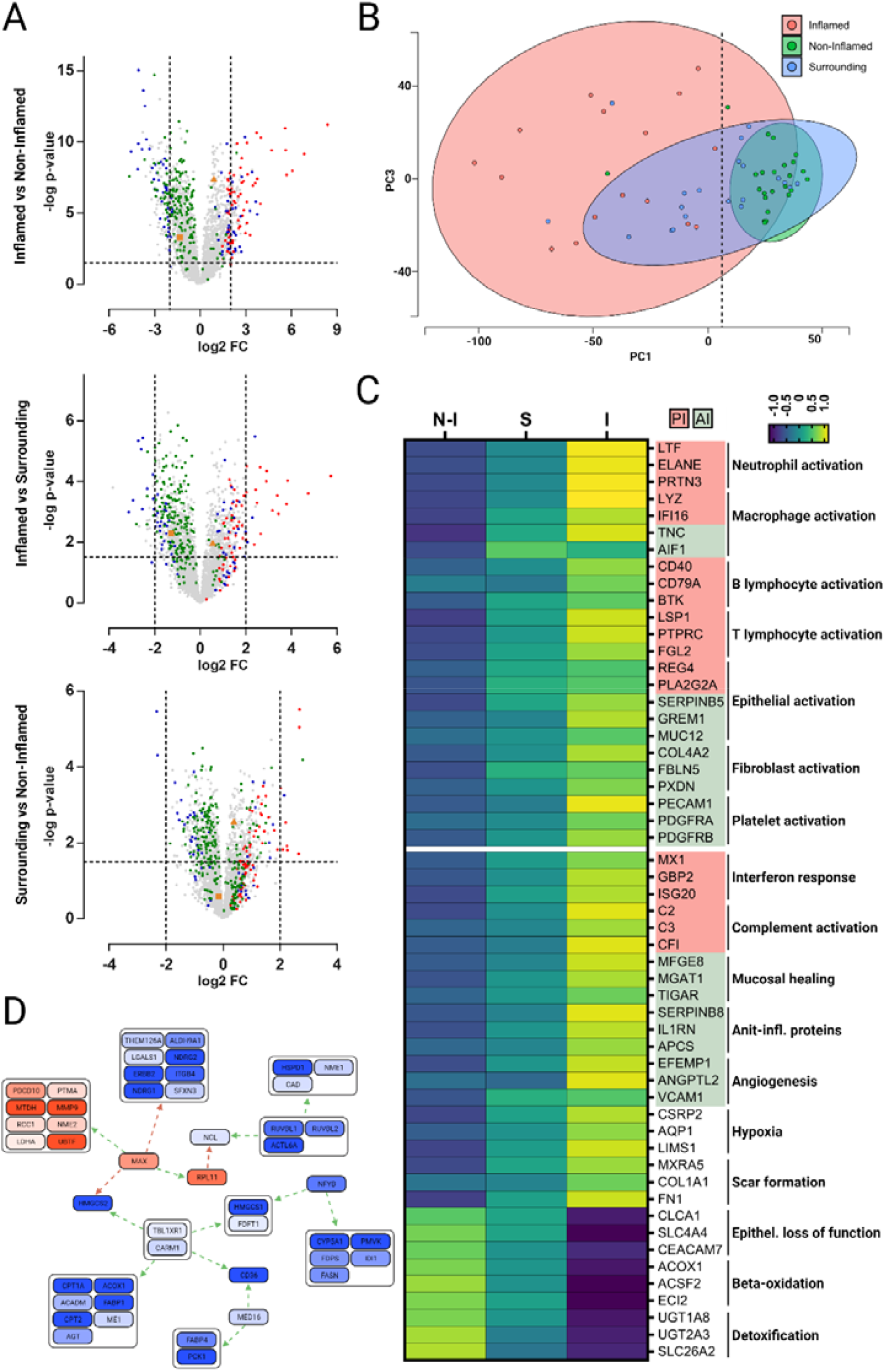
Tissue proteomics identifies pathomechanisms characteristic for UC. **(A)** Volcano plots illustrating comparisons of differentially abundant proteins in tissue samples according to histological graded categories. Proteins highlighted correspond to neutrophil granulocytes/macrophages (red), epithelial/fibroblast cells (blue) and mitochondria (green). VIM and KRT18 are highlighted as orange triangle or square, respectively. Axis were scaled according to extend of regulation, with anchor points as dotted lines at -log p-value of 1.5 (horizontal) and log2 fold change of −2 and 2 (vertical). **(B)** Principal component analysis of tissue proteomics dataset, each point representing a single colon biopsy. PC1 is plotted against PC3, allowing a complete separation of non-inflamed sections from inflamed sections (dotted line), with surrounding sections in between. **(C)** Heatmap of selected proteins of interest involved in pathophysiological processes. Color coding of gene names with red and green relates to pro-inflammatory (PI) and anti-inflammatory (AI) properties of the given protein, respectively. LFQ intensity values were z-score normalized and averaged for inflammation related grouping. **(D)** Causal-path analysis results of comparison from inflamed vs. non-inflamed tissue. Protein modules, colored from blue (lower expression) to red (higher expression), linked via direct positive (green) or negative (red) connectors form nodes of uniformly regulated proteins which are combined in hubs. For the calculation of significant causal connected regulation events, standard parameters were employed.

The inflammation-associated upregulation of VIM accompanied by a downregulation of KRT18 (Figure 2A, Supplementary Table S1) indicated a relative increase of mesenchymal cells at the cost of parenchymal cells. This interpretation was corroborated by a dominance of neutrophil-, macrophage- and fibroblast-specific proteins among the most strongly upregulated proteins in inflamed tissue (Figure 2A, C). Several epithelial proteins known to be induced upon inflammation such as REG4, PLA2G2A and GREM1 were found upregulated. In contrast, epithelial proteins characteristic for colon functions such as mucus formation were downregulated, including CLCA1, SLC4A4 and CEACAM7, suggesting a profound functional state switch of these cells. Actually, mitochondrial proteins were found to be the protein group most consistently downregulated, which is compatible with this notion (Figure 2A, C).

Characteristic marker proteins (Figure 2C, Supplementary Table S2, Supplementary data – tissue marker proteins) strongly indicated the inflammatory activation of neutrophils (LTF, ELANE and PRTN3), macrophages (LYZ, IFI16), B lymphocytes (CD40, CD79A, BTK) and T lymphocytes (LSP1, PTPRC and FGL2). This inflammatory signature was further corroborated by a marked interferon response (MX1, GBP2, ISG20) and apparent local complement activation (C2, C3, CFI). However, platelet activation, potentially promoted by FBLN1 and CD40 and evidenced by PECAM1, PDGFRA and PDGFRB, indicated an onset of wound healing activities already during acute UC. This interpretation was supported by an increased abundance of several TGF-beta induced proteins derived from macrophages (e.g. TNC, AIF1), fibroblasts (COL4A2, FBLN5, PXDN) and epithelial cells (e.g. SERPINB5, GREM1, MUC12) in addition to markers for mucosal healing (e.g. MFGE8, MGAT1, TIGAR) and other anti-inflammatory proteins such as SERPINB8, IL1RN and APCS. Furthermore, a proteome signature pointing to hypoxia (CSRP2, AQP1, LIMS1) may be causally related to angiogenesis, indicated by EFEMP1, ANGPTL2, VCAM1, and scar formation, indicated by MXRA5, COL1A1 and FN1 (Figure 2C). The downregulation of enzymes essential for beta-oxidation such as ACOX1, ACSF2 and ECI2 may also indicate a functional state switch of tissue functions potentially resulting in a loss of energy-demanding detoxification capabilities mediated by UGT1A8, UGT2A3, SLC6A2 and others.

Investigation of the tissue proteomics data using causal path analysis (Materials and Methods) indicated a hub function of the metabolic key transcription factor MAX (Figure 2D), which has also been linked to a hypoxia-induced metabolic switch promoting glycolysis above beta-oxidation.^42^

### UC pathomechanisms also affect systemic molecular plasma profiles

Proteins, eicosanoids and metabolites were analyzed in plasma samples derived from healthy controls, patients suffering from acute colitis, as well as UC patients in remission using three different LC/MS-based analysis strategies comprising 293, 72 and 494 distinct molecules (Figure 3A, Supplementary Tables S6, S7, S8), respectively. Unsupervised PCA of identified proteins separated the remission group from the healthy group, whereas the PCA of metabolomics and eicosanoid analyses separated active disease patients from healthy controls (Figure 3B). Each kind of analysis delivered significant regulatory events as displayed in volcano plots (Figure 3C) and listed in Supplementary Tables S4-S6. The application of analytes-set analyses and Data Integration Analysis, as described in Materials and Methods, further improved the separation capabilities and suggested molecular profiles associated with the clinical phenotype (Figure 4A, B). Further unsupervised hierarchical clustering of blood-borne molecules listed several biomarker candidates and separated the patient groups quite well, with some intersection between the active colitis and the remission group (Figure 4B). Figure 4C shows a molecular network of correlated regulated analytes and sub-networks generated from an integrative analyses approach comparing patients in remission with healthy controls.

**Figure 2:**
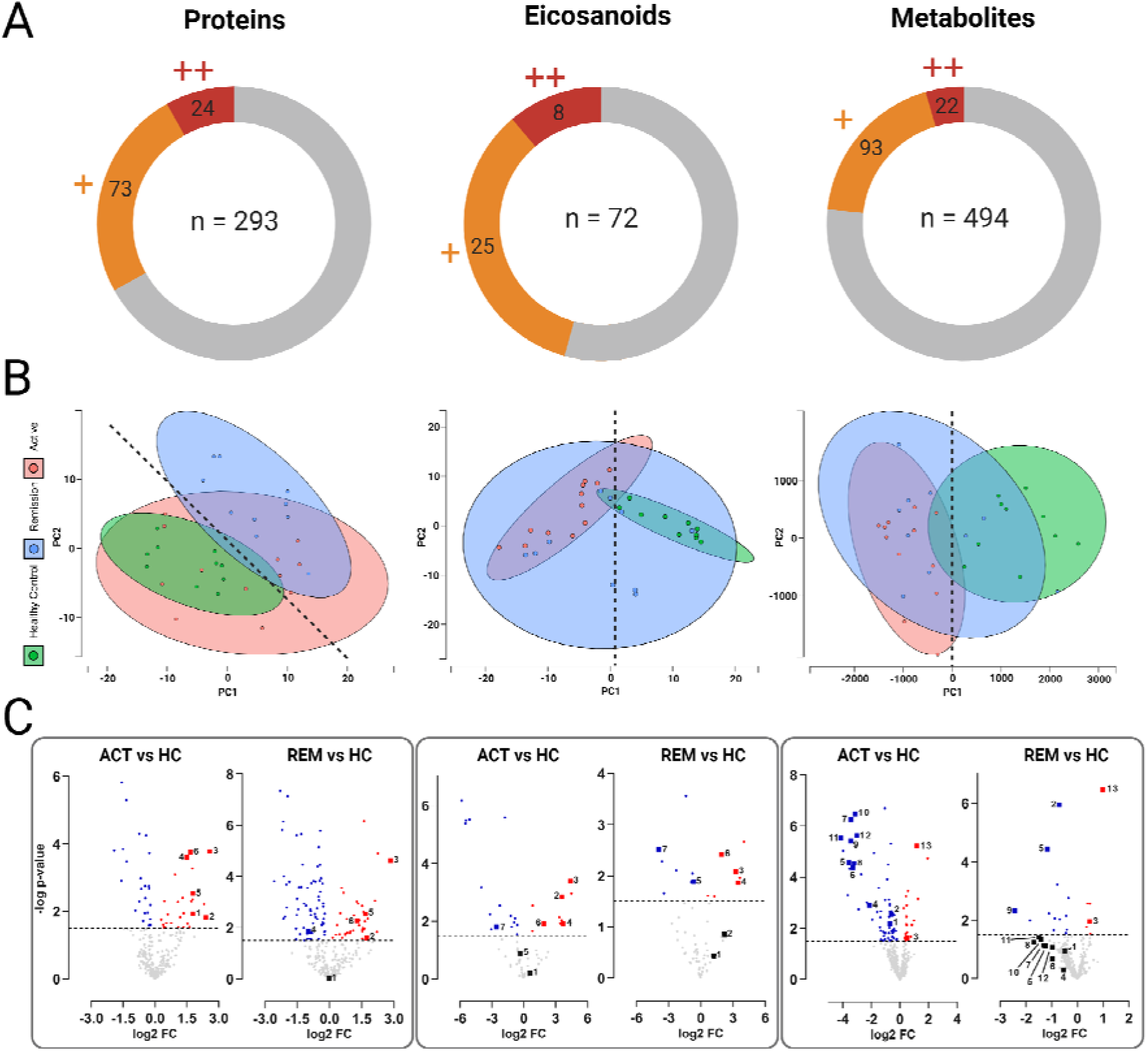
Multi-omics analysis of blood plasma. **(A)** Pie charts indicating significant regulations of proteins, eicosanoids, and metabolites derived from plasma samples. Number of experimentally determined analytes per molecule class are depicted in the middle of the respective pie chart circle. Statistical testing was performed employing a two-sided t-test (p=0.05) for the comparison between 1) UC Active versus Healthy Control and 2) UC Remission versus Healthy control. Number of analytes only significantly regulated in one of those comparisons are depicted in orange (+), analytes significantly regulated in both comparisons are depicted in red (++). **(B)** Principal component analysis of in Figure 3a illustrated datasets. Individual dots represent individual patients. 95% confidence intervals of patient cohorts are highlighted via respective coloring of areas in the plot. PCA of plasma proteins allows separation of Healthy Controls from UC Remission patients over both axis (dotted line), PCA of plasma metabolites as well as eicosanoids allowed separation of Healthy Controls from UC Active patients via PC1 (dotted lines). **(C)** Volcano plots of comparative data analysis as indicated. Significant protein regulatory events (-log p-value=1.5) between patient cohorts are highlighted in blue (down-regulation) and red (up-regulation). Selected protein candidates (left box) CRP, SAA1, ITGA2B, FBLN1, VCL and PTGDS are annotated from 1 to 6, respectively. Eicosanoid candidates (middle box) PGE2, 11-HETE, 11-HEPE, 12-HEPE, 5S-HETE, 14-HDoHE and PGD2 are annotated from 1 to 7, respectively. Metabolite candidates (right box) ornithine, aconitic acid, cystine, glycocholic acid, deoxycholic acid, glycodeoxycholic acid, glycolithocholic acid, trigonelline, p-cresol sulfate, hippuric acid, 3-indol propionic acid, indoxyl sulfate and sarcosine are annotated from 1 to 13, respectively.

**Figure 3:**
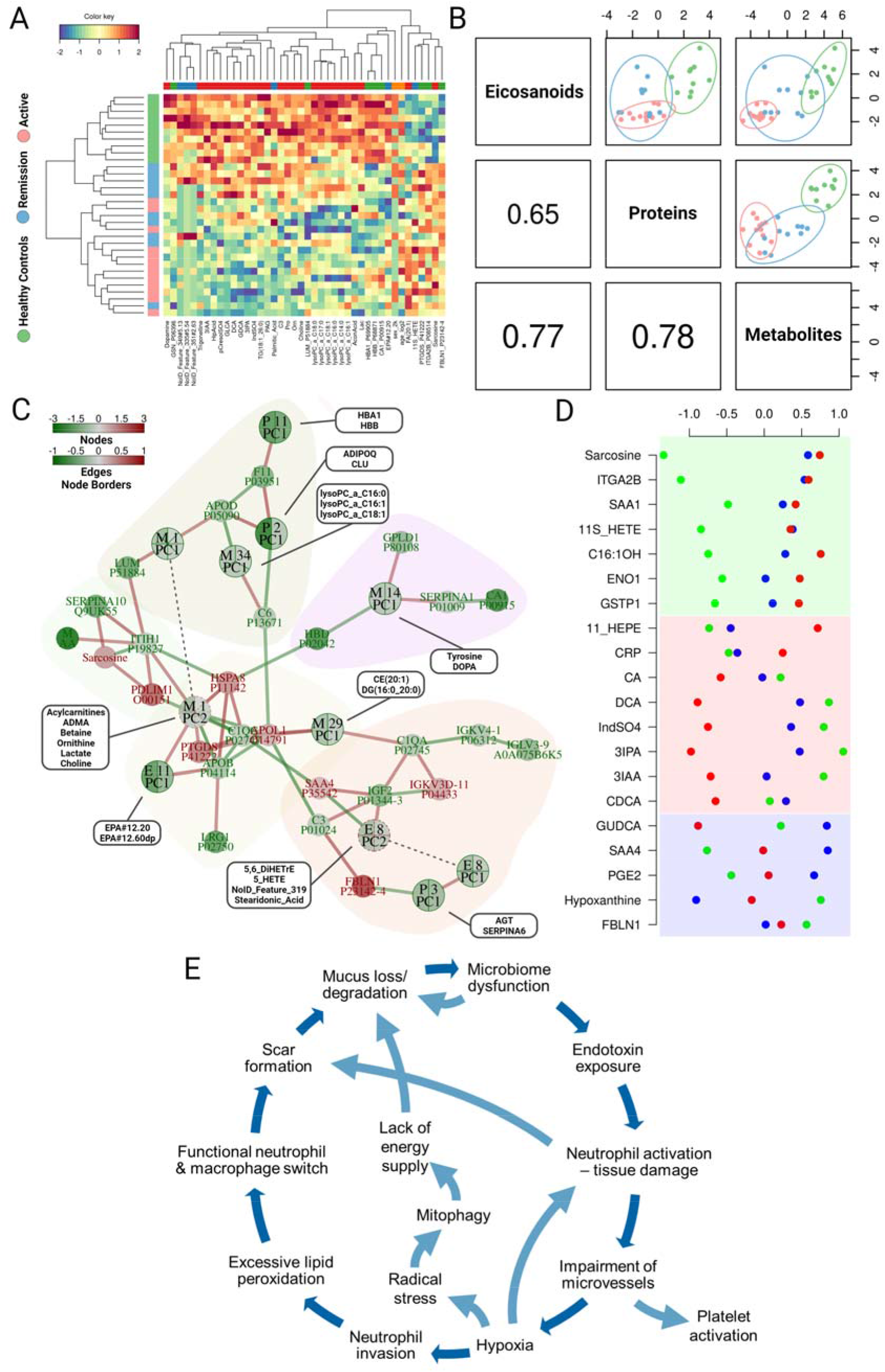
Data Integration Analysis and prognostic biomarker candidates. **(A)** Heatmap of plasma analytes (proteins, eicosanoids, and metabolites), selected by Data Integration Analysis for Biomarker discovery (DIABLO) implemented in the R-package mixOmics including information on age and sex. Eight proteins, six eicosanoids, 26 metabolites, age, and sex were selected for the first component (shown in a) and 14, six, and five, respectively, and age were selected for the second component. **(B)** Correlation plots of the first components of the DIABLO analysis using the selected analytes of each analyte type given above, color coded as in (a): green, healthy control; blue, patients in remission; and red, patients with active disease. **(C)** Network of correlated significant analytes and significant analyte sub-networks generated from an integrative analyses approach (for details see Materials and Methods) comparing patients in remission with healthy controls. Single analytes are given with their names, sub-networks of analytes are represented by their first (PC1) and sometimes second components (PC2, connected by dashed lines) and named “P” for proteins, “E” for eicosanoids, and “M” for metabolites. The second component is used if the first component represents less than 75% of the variance. The number of summarized analytes in each sub-network is indicated by the number of pie pieces in each node. The colors of the nodes and – in case of summarized sub-networks – the pie pieces of the nodes represent the log2 fold-changes between patients in remission and healthy controls, with colors in red as higher in patients with remission. If the borders of the pie pieces of the nodes representing sub-networks are in solid lines, the corresponding component is also significantly associated with the comparison of patients in remission and healthy controls. The color of the edges indicates the pairwise partial Pearson correlations between analytes and/or the Principal Component representations of the sub-networks (cut-off: |R| > 0.6). **(D)** Z-scores of analytes sorted according their behavior in the trajectory healthy control (green) - > patients in remission (blue) -> patients with acute disease (red). Green highlighted are analytes which do not normalize in patients in remission, red highlighted are analytes which get close to normal during remission, and blue highlighted are analytes which show an exceptional behavior, i.e. either the value of healthy controls is in-between the other two categories (GUDCA) or the values of healthy controls and patients in remission are at the edges and the values in patients with acute disease are in-between. **(E)** Illustration of pathophysiological mechanisms driving UC disease chronicity and proposed interconnected network depicting malicious cascades resulting in a “vicious circle”.

In an independent approach for data interpretation, molecules deregulated in plasma of UC patients were screened manually searching for potential biomarkers related to pathomechanisms evidenced by tissue proteomics. Molecules were discriminated based on significant alterations within one or two, out of two, group comparisons (active UC versus control and remission versus control, Figure 3A). Proteome profiling identified the liver-derived acute phase proteins CRP and SAA1 indicative for acute inflammation (Figure 3C). The proteins ITGA2B, FBLN1 and VCL are strongly expressed by platelets and indicate platelet involvement in UC.^43^ The detected upregulation of PTGDS, catalyzing the formation of the platelet aggregation inhibiting the eicosanoid PGD_2_^44^ and described to be upregulated upon injury,^45^ may represent a systemic response to onset of wound healing and thromboembolism – a characteristic risk for UC patients.

Eicosanoid analyses showed that the neutrophil-derived inflammation marker PGE_2_ and 5S-HETE were consistently detected but not found to be differentially regulated. The prostaglandin PGD_2_, described to be involved in the protection of the gut mucosa and promoting regeneration, was actually found downregulated.^46^ In contrast, the mainly platelet-derived lipoxygenase product 12S-HETE, the lipid peroxidation marker 11-HETE ^47^ as well as the anti-inflammatory molecules 11-HEPE, 12-HEPE and 14-HDoHE were found significantly upregulated (Figure 3C). The latter molecules were previously described to be associated with a neutrophil’s eicosanoid class switch characteristic for N2.^48^ Thus, rather anti-inflammatory contributions from platelets and neutrophils apparently dominated the UC-associated plasma eicosanoid profile.

Notably, metabolomics analyses detected the downregulation of several amino acids including ornithine. This observation may indicate reduced amino acid uptake via the gut epithelial cells in colitis patients. Increased levels of propionylcarnitine, potentially accumulating upon mitochondrial dysfunction, and a loss of aconitic acid, an essential citric acid cycle component, may relate to mitochondrial stress consistent with the notion of a mitochondrial loss (Figure 2A). Increased plasma cystine levels may relate to oxidative stress,^49^ a characteristic consequence of mitochondrial stress. The broad downregulation of the primary bile acid glycocholic acid and the secondary bile acids deoxycholic acid, glycodeoxycholic acid and glycolithocholic acid may indicate a loss of lipid synthesis or reduced lipid uptake. Various metabolites formed with the contribution of the gut microbiome such as p-cresol sulfate, hippuric acid, 3-indol propionic acid and indoxyl sulfate were found significantly decreased, indicating a loss of gut microbiome activities. Sarcosine, which has been described as oncometabolite due to its implications with hypoxia and mitochondrial stress,^50^ was found upregulated in patients with remission as well as in patients with active disease.

Several metabolic alterations may be directly related to altered levels of associated enzymes detected by tissue proteomics. Along with many other mitochondrial proteins, ACO2 was found downregulated in inflamed tissue regions, as well as cytoplasmic ACO1. This may relate to the systemic downregulation of aconitic acid mentioned above. CYP27A1, another mitochondrial enzyme presently found downregulated in inflamed tissue, catalyzes the first biosynthetic step of bile acid synthesis, and may thus relate to the above-described downregulation of bile acids. LPCAT1, an enzyme found significantly upregulated in inflamed tissue samples, was described to be upregulated upon gut microbiome dysregulation.^51^ The main substrates consumed by LPCAT1, lysophosphatidylcholines (n=14 distinct molecules) were found significantly downregulated in the plasma of colitis patients. This may promote the formation of platelet activating factors (PAFs) as described by us previously^**52**^ resulting in calcium mobilization reinforcing chronic inflammation.

### Recovery pattern of UC biomarkers upon remission indicates unresolved disease processes

Remission from acute UC was assessed with clinical parameters such as calprotectin and CRP levels in addition to several other clinical parameters (Supplementary data – UC remission patient cohort) and subjective reports regarding relief of symptoms. Only patients showing full remission were included. Remarkably, through comparison of control, remission and active UC plasma samples, we identified some candidate biomarkers not recovering back to healthy control levels, suggesting that some UC pathomechanisms may prevail after clinical remission (Figure 4D). On the one hand, the levels of bile acids CA, DCA, DCDA and IndSO_4_ and the gut microbiome-derived metabolites 3IPA and 3IAA were found close to normal values during remission, potentially indicating efficient recovery of gut functions such as lipid extraction and gut microbiome metabolism (Figure 4D, marked red). In contrast, sarcosine, the inflammation-associated molecules SAA1, 11S-HETE and the platelet-associated proteins ITGA2B and ENO1 hardly returned to normal levels, potentially indicating difficulties for the re-establishment of a normal homeostasis (Figure 4D, marked green). Indeed, several molecules involved in chronic inflammation (e.g. SAA4 and PGE2) and other pathomechanisms such as deregulated lipid uptake or hypoxia (e.g. GUDCA and hypoxanthine) were found even more deregulated upon remission when compared to the active disease states (Figure 4D, marked blue).

## Discussion

This study realized deep molecular phenotyping of UC pathomechanisms focusing on tissue proteomics and multi-omics blood plasma analyses. Tissue proteomics not only confirmed many known characteristics of UC such as neutrophil invasion and neutrophil extracellular trap as well as scar formation, but also painted a comprehensive picture of acute and chronic events.

As expected, many pro-inflammatory factors were released by tissue-resident immune cells including neutrophils, macrophages, B-cells and T-cells (Figure 2, Supplementary data). However, during inflammation, the resulting formation of oxidized lipids, together with TGF-beta and the administration of antiphlogistic drugs may initiate a functional switch of neutrophils and macrophages to a regenerative phenotype designated as N2 and M2, as characterized previously.^48, 53^ Several molecules characteristic for M2 such as TNC and AIF1 were found strongly upregulated in the acutely inflamed tissue samples, indicating the presence of polarized macrophages. Remarkably, no inflammation-associated molecules derived from fibroblasts were found upregulated in the inflamed tissue samples, whereas many TGF-beta induced proteins were found upregulated, pointing to a predominance of wound-healing activities of fibroblasts. A similar observation was made regarding epithelial cells. Several TGF-beta induced proteins such as SERPINB5 and SERPINB8, as well as proteins associated with mucosal healing such as MFGE8, MGAT1 and TIGAR (Figure 2, Supplementary data) were found upregulated in inflamed tissue samples. Furthermore, the apparent involvement of platelets also points to wound-healing activities. Thus, the present data suggested regenerative processes mainly contributed by endothelial cells, fibroblasts, platelets and epithelial cells to occur during active inflammation. This temporal coincidence represents a hallmark of chronic inflammation, suggesting that unresolved regulatory signaling mechanisms may prevail.^54^ The observed tissue regeneration efforts seem to remain somewhat unsuccessful, as the clinical phenotype shows delayed recovery and a high risk for inflammation-relapse. This raises the question, which molecular mechanisms might account for this. The following model may provide an explanation.

Recent evidence suggests that a lack of vagal nerve activities,^55^ a corresponding chronic shortage of blood supply of the guts and apparent microbiome dysregulations may contribute to ulcerative colitis initiation.^56^ Therefore, a lack of energy-demanding mucus formation in addition to mucus degradation by pathogenic microorganisms could account for increased exposure of epithelial cells to bacterial endotoxins, eventually triggering immune cell invasion. Exaggerated neutrophil activities may cause tissue damage,^57^ most notably microvascular damage further promoting a lack of oxygen supply. Epithelial cells will suffer from both, neutrophil-induced tissue destruction and a lack of oxygen required for high energy demanding activities such as mucus synthesis, transcellular transport and detoxification. Apparently, a strong angiogenic and wound-healing response is accompanying these events, potentially orchestrated by platelets activated at the surface of microvascular damage sites. Platelets are a highly plausible source for TGF-beta, apparently accounting for a large number of specific protein regulatory events currently observed in inflamed tissue samples.^58^ Several mechanisms may feed into a potential vicious cycle (Figure 4E). It is reasonable to speculate that scar formation during early provisional tissue regeneration is accompanied by a decrease in mucus formation as fibroblasts are unable to secrete mucus. More neutrophil-induced tissue damage may thus promote more scar tissue and thus less mucus formation, less detoxification capability and more vulnerability to endotoxins. Hypoxia, promoted by a lack of vagal nerve activities in addition to damaged microvessels, may further attract neutrophils,^59^ promoting an exaggerated response. An eventual functional switch to N2 may shut down 5-LOX, thus no longer promoting highly desirable phagocytotic activities of macrophages. Hypoxia may also cause mitophagy and attenuate energy-demanding processes. Continuous anti-inflammatory treatment as typically applied for UC patients may also affect stromal cells and thus interfere with regenerative mechanisms in a not yet fully understood fashion.^60^

The presently described biomarker candidates may serve as proxies for functional aberrations and thus indicate unresolved pathomechanisms in patients after remission. Successful treatment seems to recover major primary gut functions, as suggested by the corresponding biomarker levels in patients after remission. The apparent normalization of the acute inflammation marker CRP and the eicosanoid 11-HEPE supports this interpretation (Figure 4D). However, the chronic inflammation marker SAA1, the platelet activation markers ITGA2B and ENO1 and the radical stress marker 11S-HETE remain deregulated. Sarcosine, also belonging to this group, may relate to a still attenuated mitochondrial metabolism.

It seems to us that a prevalence of innate immune system-derived pathogen defense mechanisms above tissue regeneration may generally promote chronic diseases. Similar observations were reported by us with regard to chemical hepatocarcinogenesis promoted by inadequately activated Kupffer cells,^61^ and a lack of cartilage regeneration due to apparent pathogen defense activities of neutrophils.^62^

In summary, the application of a multi-omics analysis strategy allowed us to relate systemic blood-borne proteins, metabolites and lipids to local UC pathomechanisms with potential synergistic prognostic power. Prospective clinical studies are needed to test whether the present biomarker candidates will be able to fulfill our expectation regarding prognostic power supporting a more individualized colitis treatment strategy.

## Data Availability

The mass spectrometry proteomics data comprising both tissue and plasma analyses have been deposited to the ProteomeXchange Consortium (http://proteomecentral.proteomexchange.org) via the PRIDE partner repository (PMID: 30395289) with the dataset identifier PXD030775.

## Supplementary information

### Supplementary Materials and Methods

#### Supplementary data

1. Supplementary Figure S1: Age and gender distribution of UC patients.
2. Supplementary Figure S2: Histologic assessment of colon tissue samples.
3. Justification of marker proteins identified by tissue proteomics
4. Visualization of clinical parameters of UC patients

**Supplementary Table S1**: Clinical data of UC patient cohort

**Supplementary Table S2**: Clinical data of UC remission cohort

**Supplementary Table S3**: Colon tissue proteome analysis results listing LFQ values

**Supplementary Table S4**: Colon tissue proteome analysis volcano data matrices for 1-inflamed versus non-inflamed; 2-inflamed versus surrounding, 3-surrounding versus non-inflamed.

**Supplementary Table S5**: z-scores for heatmap in Figure 2

**Supplementary Table S6**: Plasma proteome analysis results listing LFQ values

**Supplementary Table S7**: Plasma eicosanoid analysis results listing nAUC values

**Supplementary Table S8**: Plasma metabolome analysis results listing μM concentration values

## Author contributions

Conceptualization: LJ, DS, NP, TF, CG

Methodology: NP, TF, CG

Investigation: LJ, DS, PB, GH, JB, SMMM, JCM, AB, BH, BW

Data processing and interpretation: LJ, DS, SMMM, AS, AB, GDF, DP, TP, CG

Visualization: LJ, PB, GH

Project administration: DS, CM, NP, CG

Supervision: TF, CG

Writing – original draft: LJ, CG

Writing – review & editing: LJ, DS, AB, SMMM, LU, CG

## Acknowledgements

This work was supported by the University of Vienna and by the Joint Metabolome Facility (University of Vienna, Medical University of Vienna), member of the VLSI (Vienna Life Science Instruments).

