## Supplementary Data for "Multi-omics empowered deep phenotyping of ulcerative colitis"

**1.)**
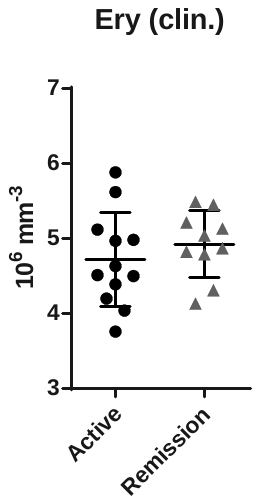

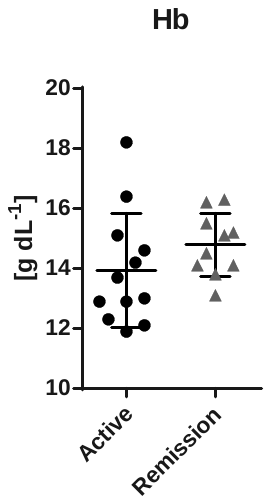

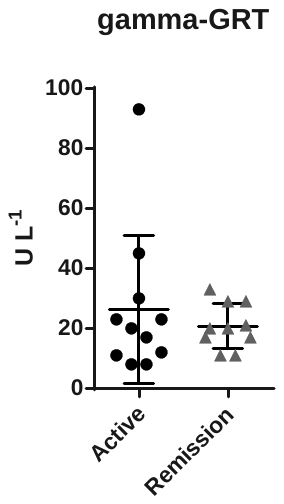
**Supplementary Figure S1**: Age and gender distribution of UC patients

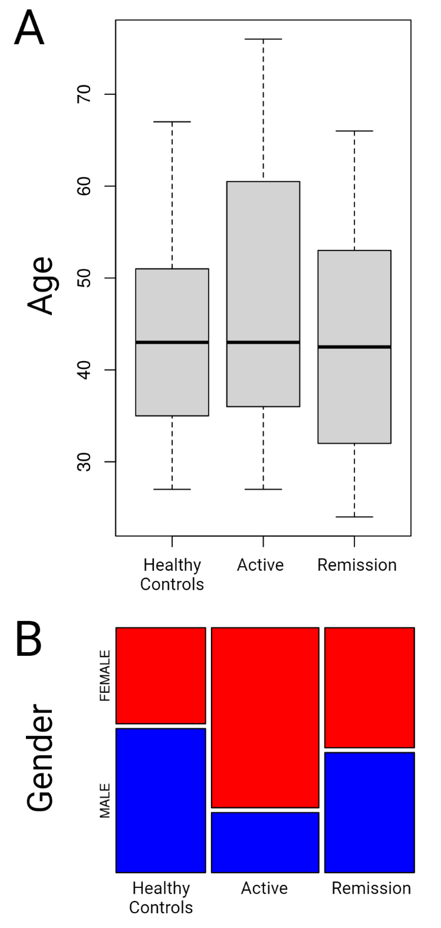

**2.) Supplementary Figure S2**

Histologic assessment of colon tissue samples. A) Colon sigmoideum, upper arrow indicates areas of higher density of inflammatory cells, lower area indicates distorted crypts with with minimal invasion of neutrophils. B) Rectum, signs of crypt elongation, arrow indicates crypt abscesses, high density of inflammatory cells in the lamina propria and a decrease in goblet cell count. C) Colon descendens, severely distorted crypts, crypt-invading neutrophils and lymphocytes present, crypt abscesses present. D) Colon sigmoideum, normal crypt shape with signs of branching, neutrophil invasion into crypts present, center of the image shows high density of inflammatory cells including eosinophils.

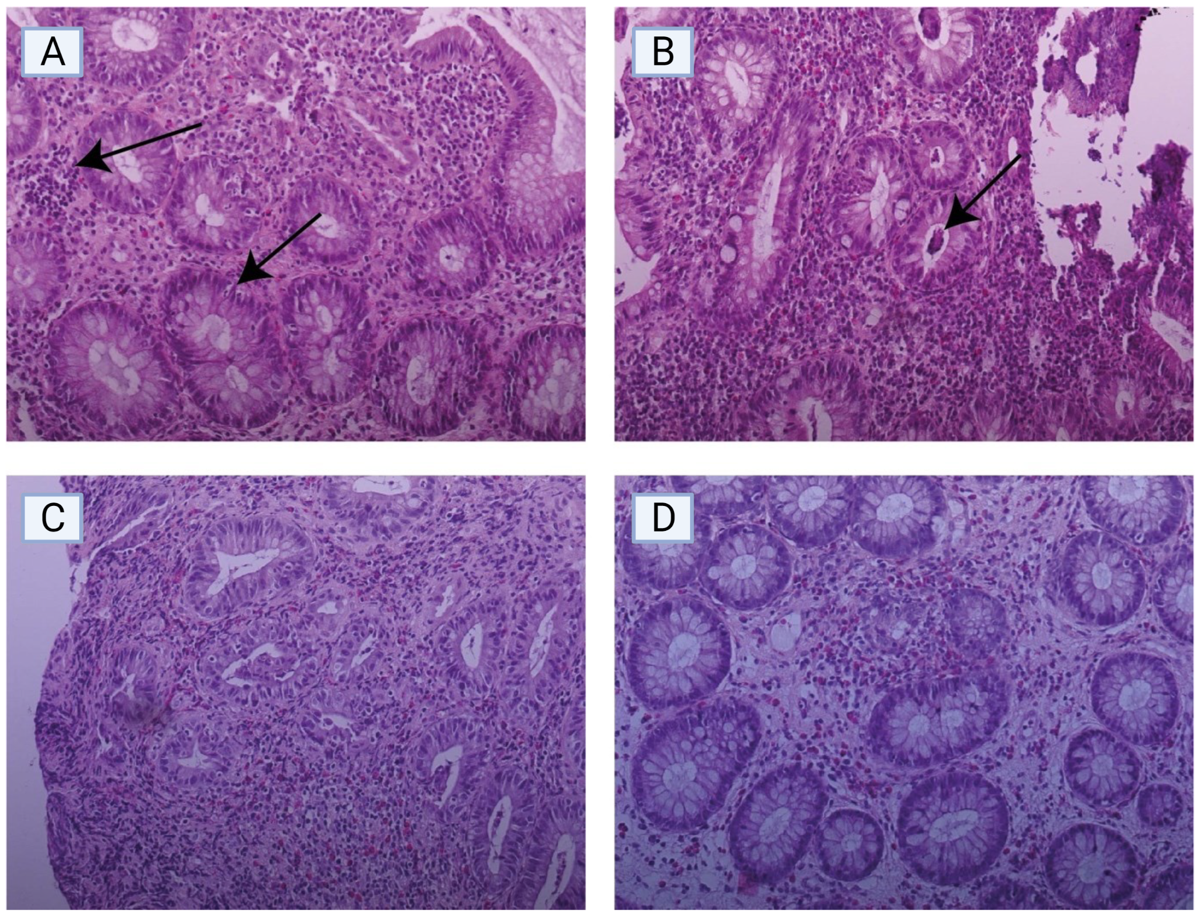

**3.) Justification of marker proteins identified by tissue proteomics**

**Neutrophil activation:**

LTF: Lactotransferrin. LTF binds to the bacterial surface and is crucial for the bactericidal functions.^1^ LTF is released by neutrophils upon neutrophil extracellular trap formation.^2^

ELANE: Neutrophil elastase. ELANE modifies the functions of natural killer cells, monocytes and granulocytes.^1^ LTF is released by neutrophils upon neutrophil extracellular trap formation.^2^ Accounting for its role in neutrophil mediated antibacterial defense, ELANE was described as potential biomarker for sepsis.^3^

PRTN3: Myeloblastin. PRTN3 is a serine protease that degrades elastin, fibronectin, laminin, vitronectin, and collagen types I, III, and IV (in vitro).^1^ Accounting for its role in neutrophil mediated antibacterial defense, PRTN3 was described as potential biomarker for sepsis.^3^

**Macrophage activation pro-inflammatory/M1:**

LYZ: Lysozyme. Lysozymes have primarily a bacteriolytic function.^1^ Accounting for its role in neutrophil mediated antibacterial defense, ELANE was described as potential biomarker for sepsis.^3^

IFI16: Gamma-interferon-inducible protein 16. Binds double-stranded DNA. Binds preferentially to supercoiled DNA and cruciform DNA structures. Seems to be involved in transcriptional regulation. May function as a transcriptional repressor. Could have a role in the regulation of hematopoietic differentiation through activation of unknown target genes.^1^ IFI16 is an inflammasome component required for DNA sensing in human macrophages.^4^

**Macrophage activation regenerative/M2:**

TNC: Tenascin. Extracellular matrix protein implicated in guidance of migrating neurons as well as axons during development, synaptic plasticity as well as neuronal regeneration.^1^ TNC produced by oxLDL-stimulated macrophages increases foam cell formation through TLR4 and scavenger receptor CD36.^5^ TNC has also been described to modulate M1/M2-macrophage polarization.^6^

AIF1: Allograft inflammatory factor 1. Actin-binding protein that enhances membrane ruffling and RAC activation. May play a role in macrophage activation and function. Promotes the proliferation of vascular smooth muscle cells and of T-lymphocytes. Enhances lymphocyte migration. Plays a role in vascular inflammation.^1^ AIF1 has been demonstrated to work as immunohistochemical marker for macrophages in multiple tissues.^7^ Importantly, AIF1 has been described to be overexpressed in M2-polarized macrophages.^8^

**B lymphocyte activation**

CD40: Transduces TRAF6- and MAP3K8-mediated signals that activate ERK in macrophages and B cells, leading to induction of immunoglobulin secretion.^1^ CD40 is an activation marker for both monocytes and B cells.^9^

CD79A: Required in cooperation with CD79B for initiation of the signal transduction cascade activated by binding of antigen to the B-cell antigen receptor complex (BCR) which leads to internalization of the complex, trafficking to late endosomes and antigen presentation. Also required for BCR surface expression and for efficient differentiation of pro- and pre-B-cells.^1^ CD79A mediated signaling is required for B cell maturation, and CD79A is used as a characteristic marker for B cells.^10^

BTK: Tyrosine-protein kinase. Non-receptor tyrosine kinase indispensable for B lymphocyte development, differentiation and signaling. Binding of antigen to the B-cell antigen receptor (BCR) triggers signaling that ultimately leads to B-cell activation.^1^

**T lymphocyte activation**

LSP1: Lymphocyte-specific protein 1. May play a role in mediating neutrophil activation and chemotaxis.^1^ LSP1 is developmentally regulated during T-cell maturation and a marker for activated T cells.^11^

PTPRC: Receptor-type tyrosine-protein phosphatase C. Protein tyrosine-protein phosphatase required for T-cell activation through the antigen receptor. Acts as a positive regulator of T-cell coactivation upon binding to DPP4.^1^

FGL2: Fibroleukin. May play a role in physiologic lymphocyte functions at mucosal sites.^1^ FGL2 is a soluble T follicular regulatory cell effector molecule binding to B cells and regulating important checkpoint molecules.^12^

**Epithelial activation/pro-inflammatory**

REG4: Regenerating islet-derived protein 4. Calcium-independent lectin displaying mannose-binding specificity and able to maintain carbohydrate recognition activity in an acidic environment. May be involved in inflammatory and metaplastic responses of the gastrointestinal epithelium.^1^ Colonic recovery from colitis-like injury was suggested to depend on Wnt-signalling affecting colonic Reg4+ epithelial cell differentiation.^13^

PLA2G2A: Phospholipase A2, membrane associated. Secretory calcium-dependent phospholipase A2 that primarily targets extracellular phospholipids with implications in host antimicrobial defense, inflammatory response and tissue regeneration.^14^ PLA2G2A was described in mouse IBD models to be associated with increased colitis susceptibility.^15^

**Epithelial activation/regenerative**

SERPINB5: Serpin B5. Tumor suppressor. It blocks the growth, invasion, and metastatic properties of mammary tumors. As it does not undergo the S (stressed) to R (relaxed) conformational transition characteristic of active serpins, it exhibits no serine protease inhibitory activity.^1^ SERPINB5 was already demonstrated to be deregulated in ulcerative colitis and colon carcinoma.^16, 17^

GREM1: Gremlin-1. Cytokine that may play an important role during carcinogenesis and metanephric kidney organogenesis, as a BMP antagonist required for early limb outgrowth and patterning in maintaining the FGF4-SHH feedback loop.^1^ GREM1 upregulation upon inflammatory bowel disease was demonstrated to be required for adaptive reprogramming of intestinal regeneration. ^18^

MUC12: Mucin-12. Involved in epithelial cell protection, adhesion modulation, and signaling. May be involved in epithelial cell growth regulation. Stimulated by both cytokine TNF-alpha and TGF-beta in intestinal epithelium.^19^

**Fibroblast activation**

COL4A2: Collagen alpha-2(IV) chain. Type IV collagen is the major structural component of glomerular basement membranes (GBM), forming a 'chicken-wire' meshwork together with laminins, proteoglycans and entactin/nidogen.^1^ A role of α1 and α2 chains of type IV collagen in early fibrotic lesions has been demonstrated.^20^

FBLN5: Fibulin-5. Essential for elastic fiber formation, is involved in the assembly of continuous elastin (ELN) polymer and promotes the interaction of microfibrils and ELN in a three-dimensional fibroblast model.^21^ Was found upregulated in fibroblasts upon treatment with TGF-beta.^22^

PXDN: Peroxidasin homolog. Displays low peroxidase activity and is likely to participate in H_2_O_2_ metabolism and peroxidative reactions in the cardiovascular system. Plays a role in extracellular matrix formation.^23^ Contributes to fibrosis progression through extensive collagen cross-linking.^24^

**Platelet activation**

PECAM1: Platelet endothelial cell adhesion molecule. Cell adhesion molecule which is required for leukocyte transendothelial migration (TEM) under most inflammatory conditions. ^25^

PDGFRA: Platelet-derived growth factor receptor alpha. Tyrosine-protein kinase that acts as a cell-surface receptor for PDGFA, PDGFB and PDGFC and plays an essential role in the regulation of embryonic development, cell proliferation, survival and chemotaxis.^1^

PDGFRB: Platelet-derived growth factor receptor beta. Tyrosine-protein kinase that acts as a cell-surface receptor for PDGFA, PDGFB and PDGFC and plays an essential role in the regulation of embryonic development, cell proliferation, survival and chemotaxis.^1^

**Interferon response**

MX1: Interferon-induced GTP-binding protein Mx1. Interferon-induced dynamin-like GTPase with antiviral activity against a wide range of RNA viruses and some DNA viruses.^1^ Strongly upregulated upon inflammatory stimulation in human peripheral blood mononuclear cells.^26^

GBP2: Guanylate-binding protein 2. Exhibits antiviral activity against influenza virus. Promotes oxidative killing and delivers antimicrobial peptides to autophagolysosomes, providing broad host protection against different pathogen classes.^1^ GBP2 may indicate an efficient T cell response.^27^

ISG20: Interferon-stimulated gene 20 kDa protein. Interferon-induced antiviral exoribonuclease that acts on single-stranded RNA and also has minor activity towards single-stranded DNA. Exhibits antiviral activity against RNA viruses including hepatitis C virus (HCV), hepatitis A virus (HAV) and yellow fever virus (YFV) in an exonuclease-dependent manner.^28^

**Complement activation**

C2: Complement C2. Component C2 which is part of the classical pathway of the complement system is cleaved by activated factor C1 into two fragments: C2b and C2a. C2a, a serine protease, then combines with complement factor C4b to generate the C3 or C5 convertase.^1^

C3: Complement C3. C3 plays a central role in the activation of the complement system. Its processing by C3 convertase is the central reaction in both classical and alternative complement pathways. After activation C3b can bind covalently, via its reactive thioester, to cell surface carbohydrates or immune aggregates.^1^

CFI: Complement factor I. Trypsin-like serine protease that plays an essential role in regulating the immune response by controlling all complement pathways. Inhibits these pathways by cleaving three peptide bonds in the alpha-chain of C3b and two bonds in the alpha-chain of C4b thereby inactivating these proteins.^29^

**Mucosal healing**

MFGE8: Lactadherin. Plays an important role in the maintenance of intestinal epithelial homeostasis and the promotion of mucosal healing.^30^

MGAT1: Alpha-1,3-mannosyl-glycoprotein 2-beta-N-acetylglucosaminyltransferase. Initiates complex N-linked carbohydrate formation. Essential for the conversion of high-mannose to hybrid and complex N-glycans. MGAT1 is a characteristic transcriptional target of Wnt/β-catenin signaling pathway,^31^ which is activated upon wounding and regulates important steps of tissue regeneration.^32^

TIGAR: Fructose-2,6-bisphosphatase TIGAR. Acts as a negative regulator of glycolysis by lowering intracellular levels of fructose-2,6-bisphosphate in a p53/TP53-dependent manner, resulting in the pentose phosphate pathway (PPP) activation and NADPH production.^33^ TIGAR has been demonstrated to be required for efficient intestinal tissue regeneration.^34^

**Anti-inflammatory proteins**

SERPINB8: Serpin B8. Has an important role in epithelial desmosome-mediated cell-cell adhesion.^1^ The intracellular serpin, proteinase inhibitor 8 (PI8/Serpinb8), can inhibit furin, a prohormone convertase involved in inflammation, prohormone processing and extracellular matrix remodeling and may thus contribute to tissue repair.^35^

IL1RN: Interleukin-1 receptor antagonist protein. Inhibits the activity of interleukin-1 by binding to receptor IL1R1 and preventing its association with the coreceptor IL1RAP for signaling.^1^ IL1RN has been demonstrated to represent a physiologically relevant way to establish an anti-inflammatory environment.^36^

APCS: Serum amyloid P-component. Can interact with DNA and histones and may scavenge nuclear material released from damaged circulating cells.^1^ APCS has been demonstrated to attenuate inflammation and inflammation-associated tissue damage.^37^

**Angiogenesis**

EFEMP1: EGF-containing fibulin-like extracellular matrix protein 1. Binds EGFR, the EGF receptor, inducing EGFR autophosphorylation and the activation of downstream signalling pathways. May play a role in cell adhesion and migration.^1^ EFEMP1 expression has been demonstrated to promote angiogenesis.^38^

ANGPTL2: Angiopoietin-related protein 2. Induces sprouting in endothelial cells through an autocrine and paracrine action.^1^ The hypoxia-inducible gene ANGPTL2 has been demonstrated to facilitate tumor proliferation, metastasis and angiogenesis in osteosarcoma.^39^

VCAM1: Vascular cell adhesion protein 1. Important in cell-cell recognition. Appears to function in leukocyte-endothelial cell adhesion. Interacts with integrin alpha-4/beta-1 (ITGA4/ITGB1) on leukocytes, and mediates both adhesion and signal transduction.^1^ The potential tumor-promoting role of VCAM1 was attributed to its functional relevance for tumor growth, metastasis, angiogenesis.^40^

**Hypoxia**

CSRP2: Cysteine and glycine-rich protein 2. Drastically downregulated in response to PDGF-BB or cell injury, that promote smooth muscle cell proliferation and dedifferentiation.^1^ The expression of CSRP2 has been demonstrated to be promoted by hypoxia.^41^

AQP1: Aquaporin-1. Forms a water-specific channel that provides the plasma membranes of red cells and kidney proximal tubules with high permeability to water, thereby permitting water to move in the direction of an osmotic gradient.^1^ AQP1 has been demonstrated to be upregulated by hypoxia in various diseases such as neuroblastoma.^42^

LIMS1: LIM and senescent cell antigen-like-containing domain protein 1. Adapter protein in a cytoplasmic complex linking beta-integrins to the actin cytoskeleton, bridges the complex to cell surface receptor tyrosine kinases and growth factor receptors. Involved in the regulation of cell survival, cell proliferation and cell differentiation.^1^ LIMS1 has been demonstrated to be induced upon hypoxia and to be crucial for tumor adaptation to oxygen-glucose deprivation conditions.^43^

**TGF-beta target genes/ scar formation**

MXRA5: Matrix-remodeling-associated protein 5. In kidney, has anti-inflammatory and anti-fibrotic properties by limiting the induction of chemokines, fibronectin and collagen expression in response to TGB1 and pro-inflammatory stimuli.^1^ MXRA5 expression is induced by TGF-beta and regulates inflammatory processes and fibrosis.^44^

COL1A1: Collagen alpha-1(I) chain. Type I collagen is a member of group I collagen (fibrillar forming collagen).^1^ Persistent TGF-beta 1 function has been demonstrated to cause excessive fibrosis and COL1A1 formation and ultimately scarring of both skin and internal organs.^45^

FN1: Fibronectin. Fibronectins are involved in cell adhesion, cell motility, opsonization, wound healing, and maintenance of cell shape.^1^ The role of fibronectin, collagen I and other ECM proteins in fibroblast-mediated scar formation has been described in detail.^46^

**Epithelial loss of function**

CLCA1: Calcium-activated chloride channel regulator 1. May be involved in mediating calcium-activated chloride conductance. May play critical roles in goblet cell metaplasia, mucus hypersecretion, cystic fibrosis and AHR. May be involved in the regulation of mucus production and/or secretion by goblet cells. Involved in the regulation of tissue inflammation in the innate immune response.^1, 47^

SLC4A4: Electrogenic sodium bicarbonate cotransporter 1. Electrogenic sodium/bicarbonate cotransporter with a Na^+^:HCO3^-^ stoichiometry varying from 1:2 to 1:3. May regulate bicarbonate influx/efflux at the basolateral membrane of cells and regulate intracellular pH.^1^ Hypoxia is also able to induce the expression of SLC4A4 affecting cell growth and migration.^48^

CEACAM7: Carcinoembryonic antigen-related cell adhesion molecule 7. CEACAM7 is specifically expressed at the apical surface of highly differentiated epithelial cells in the colorectal mucosa. Downregulation of CEACAM7 and upregulation of CEACAM6 expression in hyperplastic polyps and early adenomas represent some of the earliest observable molecular events leading to colorectal tumors.^49^

**Beta oxidation**

ACOX1: Peroxisomal acyl-coenzyme A oxidase 1. Catalyzes the desaturation of acyl-CoAs to 2-trans-enoyl-CoAs. First enzyme of the fatty acid beta-oxidation pathway.^1^

ACSF2: Medium-chain acyl-CoA ligase ACSF2, mitochondrial. Acyl-CoA synthases catalyze the initial reaction in fatty acid metabolism, by forming a thioester with CoA.^50^

ECI2: Enoyl-CoA delta isomerase 2. Able to isomerize both 3-cis and 3-trans double bonds into the 2-trans form in a range of enoyl-CoA species.^1, 51^

**Detoxification**

UGT1A8: UDP-glucuronosyltransferase 1A8. UDP-glucuronosyltransferase (UGT) that catalyzes phase II biotransformation reactions in which lipophilic substrates are conjugated with glucuronic acid to increase the metabolite's water solubility, thereby facilitating excretion into either the urine or bile. 1 Essential for the elimination and detoxification of drugs, xenobiotics, carcinogens, and endogenous compounds.^1, 52^

UGT2A3: UDP-glucuronosyltransferase 2A3. UDP-glucuronosyltransferases catalyze phase II biotransformation reactions in which lipophilic substrates are conjugated with glucuronic acid to increase water solubility and enhance excretion. They are of major importance in the conjugation and subsequent elimination of potentially toxic xenobiotics and endogenous compounds.^1^

SLC26A2: Sulfate transporter. Downregulation of this organic solute carrier was found characteristic for Crohn´s disease.^53^

**4.) Visualization of clinical parameters of UC patients**

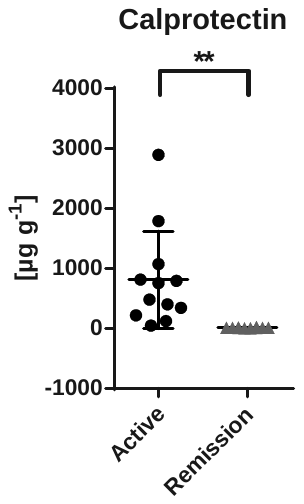

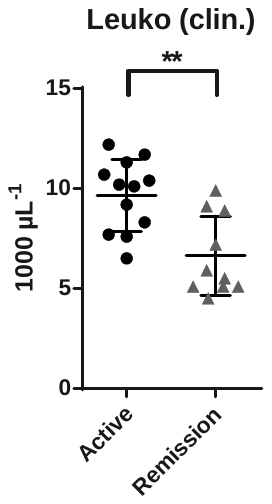

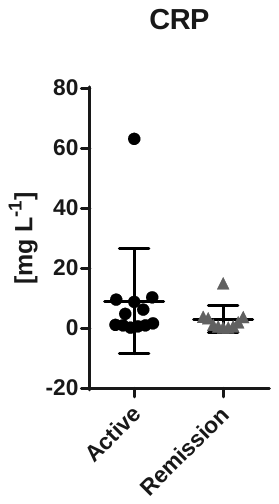

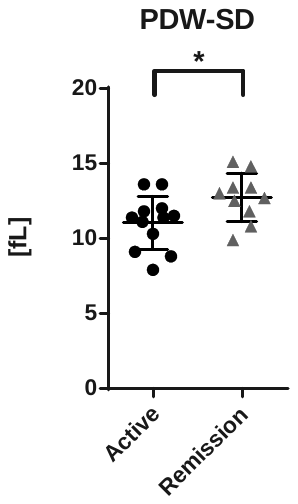

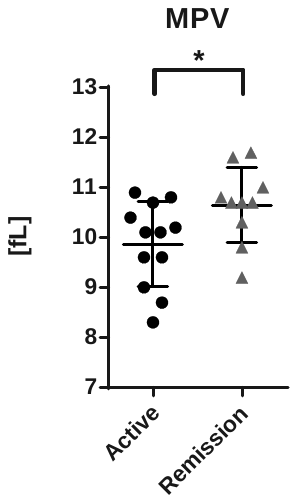

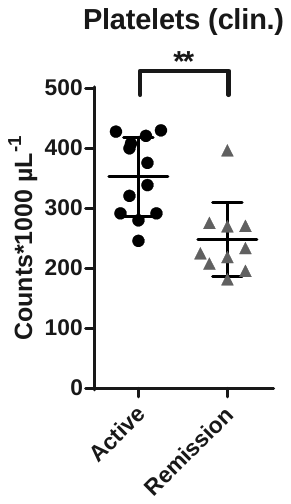

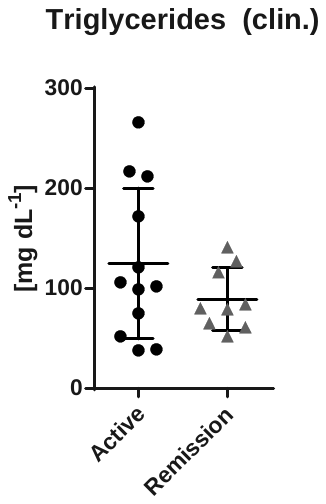

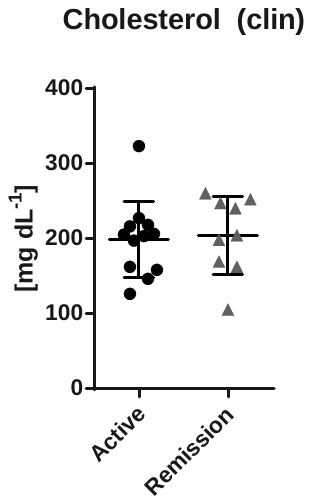

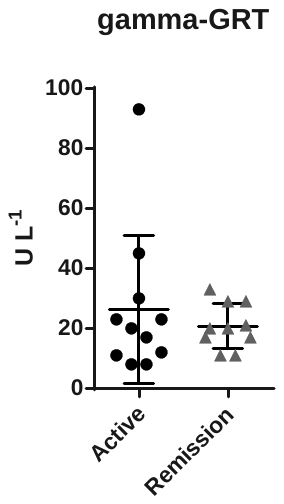

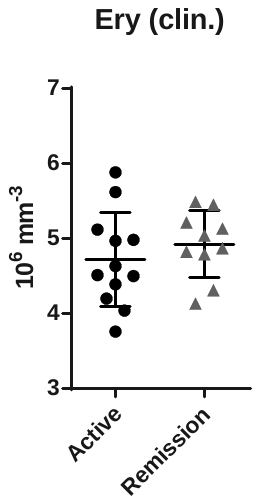

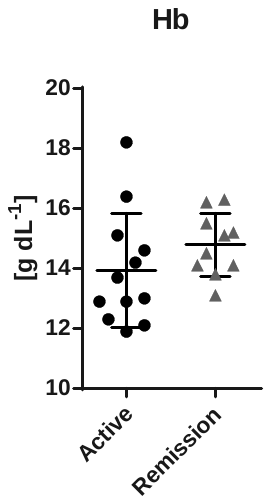

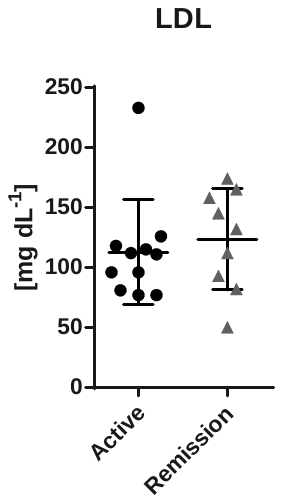

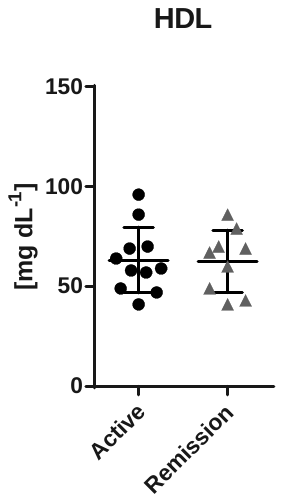
