## Supplementary_Materials_Methods for "Multi-omics empowered deep phenotyping of ulcerative colitis"

**Supplementary Material and Methods**

**Processing of plasma and tissue samples**

During routine colonoscopy, biopsies of rectum, colon sigmoideum, colon descendens, colon transversum and colon ascendens were split into two parts. The biopsy part designated for proteomic analysis was washed thoroughly with ringer’s solution, placed in a cryo-tube and stored at -80°C. Blood plasma was collected by centrifugation of full blood mixed with EDTA at 1500g for 10 minutes and subsequently stored at -80°C.

**Plasma and tissue proteomics**

Lysis buffer (8M Urea, 50mM TEAB, 5% SDS) was added to EDTA plasma at a ratio of 1:10 and to tissue samples (100µL) which were lysed via ultrasonic rod. Protein concentrations were determined via BCA-assay and 20µg of protein were used for further processing. For protein digestion, a ProtiFi S-trap^TM^ protocol was employed as described previously.^1^ In short, proteins were reduced and alkylated using DTT and IAA, respectively, followed by addition of trapping buffer. Samples were loaded onto S-trap^TM^ cartridges and digested with Trypsin/Lys-C for 2h at 37°C. Supernatants containing the peptides were eluted, dried, reconstituted in 5µL formic acid containing 10fmol of 4 synthetic standard peptides and diluted with 40µL mobile phase A. LC-MS/MS measurements were performed on a timsTOF Pro mass spectrometer (Bruker Daltonics) hyphenated with a Dionex UltiMate^TM^ 3000 RSLCnano system (Thermo Scientific). Parameters for analyses were an adapted version of a previously published method.^1^ 0.5µL and 5µL of plasma and tissue proteomic samples, respectively, were loaded onto an Acclaim^TM^ PepMap^TM^ C18 100 pre-column (Thermo Fisher Scientific) at a flow rate of 10µL/min using mobile phase A and eluted onto an Aurora Series emitter column (Ionopticks) applying a flow rate of 300nL/min. Separation was achieved by applying a gradient of 8% to 40% mobile phase B over 55min and 90min for plasma and tissue samples, respectively. Data analysis was performed using MaxQuant (version 1.6.17.0) employing the Andromeda search engine for protein identification against the UniProt database (12/2019, 20380 entries).^2^ Search parameters were set as previously described.^1^ Fixed modifications included carbamidomethylation of cysteine and methionine oxidation, N-terminal protein acetylation was set as variable modification. For the data evaluation in Perseus, data was grouped and filtered according to missing values. A total of 5 or 3 valid values had to be present in at least one cohort in tissue or plasma samples, respectively, to be considered a valid identification. Data was log2 transformed and imputation was performed according normal distribution with a down-shift of 1.8 sigma and a width of 0.3 sigma. For the group wise comparison, two-sided t-tests were performed.

**Eicosanoid analysis**

EDTA plasma (0.4 mL; freshly thawed on ice) was added to 1.6 mL cold ethanol (EtOH, abs. 99%, -20°C; AustroAlco) containing 100 nM of each internal standard (12S-HETE-d8, 15S-HETE-d8, 5-Oxo-ETE-d7, 11.12-DiHETrE-d11, PGE-d4, 20-HETE-d6; Cayman Europe) and kept overnight at -20°C to allow for protein precipitation. The suspension was centrifuged for 30 min at 4°C (4536 g, 9 DCC, 7 ACC) and the supernatant was transferred to a new 15 ml FalconTM tube. Afterwards, ethanol was evaporated via vacuum centrifugation at 37°C until the original sample volume was restored. Samples were loaded on preconditioned StrataX solid phase extraction (SPE) columns (30 mg mL-1; Phenomenex) using Pasteur pipettes, washed with 5 mL MS grade water and elution of eicosanoids was achieved with 500 µL ice cold methanol (MeOH abs.; VWR International) containing 2% formic acid (FA; Sigma-Aldrich). Methanol was evaporated under a gentle N2 stream at room temperature and dried samples were reconstituted in 150 µL reconstitution buffer (H2O/ACN/MeOH + 0.2%FA - 65:31.5:3.5), including another set of 10-100 nM internal standards (5S-HETE-d8, 14.15-DiHETrE-d11, 8-iso-PGF2a-d4; Cayman Europe, Tallinn, Estonia). Eicosanoids were analyzed with a Thermo ScientificTM VanquishTM (UHPLC) system coupled to a Q ExactiveTM HF Quadrupole-OrbitrapTM mass spectrometer (Thermo Fisher Scientific, Austria), equipped with a HESI source for negative ionization. In short, eicosanoids were separated on a Kinetex® C18-column (2.6 μm C18 100 Å, LC Column 150 x 2.1 mm; Phenomenex®) at a flow rate of 200 µL min-1. Per injection, 20 µL of the sample were loaded and all samples were analyzed in two technical replicates. The 20 min UHPLC method included a gradient flow profile (mobile phase A: H2O + 0.2% FA, mobile phase B: ACN:MeOH (90:10) + 0.2% FA) starting at 35% B increasing to 90% B (1-10 min), further going up to 99% B in 0.5 min and held for 5 min. Afterwards solvent B was decreased to a level of 35% in 0.5 min and the column was equilibrated for 4 min. The column oven temperature was set to 40°C. Mass spectrometric resolution on the MS1 level was set to 60,000 (at m/z = 200) with a scan range from 250 to 700 m/z. The two most abundant precursor ions were selected for fragmentation (HCD 24 normalized collision energy), preferentially from an inclusion list containing 31 m/z values specific for eicosanoids and their precursor molecules. The resulting fragments were analyzed on the MS2 level at a resolution of 15,000 (at m/z = 200). Operating in negative ionization mode, a spray voltage of 2.2 kV and a capillary temperature of 253°C were applied. Sheath gas was set to 46 and the auxiliary gas to ten arbitrary units. Raw files generated by the Q ExactiveTM HF Quadrupole-OrbitrapTM mass spectrometer were checked manually using Thermo XcaliburTM 4.1.31.9 (Qual browser). Spectra were compared with reference spectra from the Lipid Maps depository library from July 2018.^3^ Peaks were integrated using the TraceFinderTM software package (version 4.1 - Thermo Scientific).

**Targeted metabolomics experiments**

EDTA plasma samples (10 µL) of patients were analyzed by a targeted metabolomic assay. Targeted metabolomics experiments were conducted using the MxP® Quant 500 Kit (Biocrates Life Sciences AG), which enables the detection and (semi)quantification of up to 631 analytes, including 40 acylcarnithines, 1 alkaloid, 1 amine oxide, 50 amino acid related metabolite, 15 bile acids, 9 biogenic amines, 7 carboxylic acids, 28 ceramides, 22 cholesteryl esters, 1 cresol, 44 diacylglycerols, 8 dihydroceramides, 12 fatty acids, 90 glycerophospholipids, 34 glycosylceramides, 4 hormones, 4 indole derivatives, 2 nucleobase related metabolites, 15 sphingolipids, 242 triacylglycerols, the sum of hexoses and 1 vitamin/cofactor. A total of 494 metabolites showed signal intensities within the quantification window and were further evaluated. Measurements were carried out using LC-MS and flow injection (FIA)-MS analyses on a Sciex 6500+ series mass spectrometer coupled to an ExionLC AD chromatography system (AB Sciex), utilizing the Analyst 1.7.1 software with hotfix 1 (AB SCIEX). All required standards, quality controls and eluents were included in the kit, as well as the chromatographic column for the LC-MS/MS analysis part. Phenyl isothiocyanate (Sigma-Aldrich) was purchased separately and was used for derivatization of amino acids and biogenic amines according to the kit manual. Preparation of the measurement worklist as well as data validation and evaluation was performed with the software supplied with the kit (MetIDQ-Oxygen-DB110-3005, Biocrates Life Sciences).

**Statistical methods**

Multi-omics integration was done in GNU R following two methods, DIABLO (Data Integration Analysis for Biomarker discovery), implemented in the R-package mixOmics and an own developed method.^4^ For both approaches the single omics data types were quality controlled and prepared as follows. Protein data were imputed and log_2_ transformed as described above. All Eicosanoid values which were below the detection limit were imputed with the respective minimal value divided by the square root of two and all values were sub sequentially log_2_ transformed. Metabolite values were log_2_ transformed with an offset of 0.01. The age of patients was also log_2_ transformed.

All four datasets (proteins, eicosanoids, metabolites, and age/sex) were used together for the N-integration discriminant analysis with DIABLO implemented in the R-package mixOmics. In the design matrix the connection link of all the blocks was set to 0.2 and the number of components to be include in the model to two (restricted by the two analytes in the age/sex block). The performance of the models was evaluated by 10 times repeated 10-fold cross-validations, yielding best balanced error rates using the Mahalanobis distance measure of 0.33 for one component and 0.17 for two components, respectively. Finally, the analytes to be kept in the model was optimized by a final tuning step (cross-validation as above), setting the number of components to two and the distance measure to Mahalanobis and allowing a maximum of 10% of proteins and eicosanoids and 20% of metabolites in the model. The final model selected eight proteins, six eicosanoids, 26 metabolites and age and sex for the first component (shown in Figure 4a) and 14 proteins, six eicosanoids, five metabolites and age for the second component.

For a more fine-grained analysis following the methods described in Svoboda *et al.*,^5^ Bekos *et al.*,^4^ and Muqaku *et al.*,^6^ with the following steps were performed. For all selection steps in the following analysis a false discovery rate (FDR) cutoff of 5% was chosen.

1) For all three analyte types single analytes significantly with the differences between i) healthy controls and acute diseased patients, ii) healthy controls and patients in remission, and iii) patients in remission and acute diseased ones (*i.e.* the so called contrasts), were determined using linear modeling and an empirical Bayesian approach to moderate the standard errors across analytes, *i.e.*, shrink towards a common value, as implemented in the R-package limma.^7^ Information of age and sex was included in the models and therefore results were corrected for these two possible confounders.

2) Using graphical Gaussian modeling (GGM) implemented in R-package GGMselect^8^ for each analyte type sub-networks were identified and tested if significantly associated with the contrasts under analysis and–if yes–were used for subsequent integration over all analyte types. This step was done to reduce the number of analytes for a more clearly represented integration and easier interpretation. The principle behind GGM is to use partial correlations as a measure of independence of any two analytes which allows for distinguishing direct from indirect interactions. The tuning parameter K for the penalty function was varied between 1 and 6 in 0.5 steps and the function *selectFast* *[family = c(“C01,” “LA,”)]* was used for optimizing the model employing the C01 and the Lasso-And (LA) algorithm (https://cran.r-project.org/web/packages/GGMselect/vignettes/Notice.pdf). At each K the resulting analyte-sub-networks were used for gene set analyses (GSA, using raw p-values and regulation direction from the single analyte analyses with function *runGSA*, implemented in R-package piano) to determine the significant associations with all contrasts. Finally the K-value was chosen which yielded the largest number of contrast specific significantly associated subnetworks. All of these significantly associated subnetworks were summarized by the first (PC1) or first two principal components (PC1 and PC2; if the proportion of the explained variance of the first component was below 75%).

3) For each contrast all Principal Components for all significant subnetworks and all remaining significant single analytes over all omics type were collected together with the information of age and sex and partial correlations were calculated between all of them (R-package ppcor v1.1).^9^

4) Finally, a network was plotted for every contrast showing all significant single analytes and sub-networks (represented by the first (two) Principal Components) and their partial correlations with each other (cutoff |R| > 0.6). Color of nodes (single analytes) and pie pieces of nodes (members of each sub-network) represent the log_2_ fold-change between the two compared groups of each contrast and the color of the edges represents the R-value of the correlations between the nodes.
